# Cardiopulmonary Resuscitation Without Aortic Valve Compression Increases Chances of Return of Spontaneous Circulation for Patients Experiencing Out-of-hospital Cardiac Arrest

**DOI:** 10.1101/2023.07.17.23292797

**Authors:** Sheng-En Chu, Chun-Yen Huang, Chiao-Yin Cheng, Chun-Hsiang Chan, Hsuan-An Chen, Chin-Ho Chang, Kuang-Chau Tsai, Kuan-Ming Chiu, Matthew Huei-Ming Ma, Wen-Chu Chiang, Jen-Tang Sun

**Author notes:** **Corresponding authors:** Jen-Tang Sun, MD, MSc, Department of Emergency Medicine, Far Eastern Memorial Hospital, No.21, Sec. 2, Nan-Ya South Rd., Ban-Qiao Dist., New Taipei City, Taiwan; Wen-Chu Chiang, MD, PhD, Department of Emergency Medicine, National Taiwan University Hospital, Yun-Lin Branch, Douliu City, Taiwan, No. 7, Chung-Shan S. Rd, Taipei, Taiwan.

## Abstract

**Background:** Current cardiopulmonary resuscitation (CPR) guidelines recommend that chest compressions should be applied at “the center of the chest.” However, in approximately 50% of patients experiencing out-of-hospital cardiac arrest (OHCA) the aortic valve (AV) is reportedly compressed, potentially obstructing blood flow and worsening prognosis. We aimed to use resuscitative transesophageal echocardiography (TEE) to elucidate the impact of compressed vs. uncompressed AV on outcomes of adult patients experiencing OHCA.

**Methods:** This prospective single-center observational cohort study included patients experiencing OHCA who underwent resuscitative TEE in the emergency department (ED). Exclusion criteria were early return of spontaneous circulation (ROSC) before TEE, resuscitative endovascular balloon occlusion of the aorta (REBOA) or extracorporeal membrane oxygenation (ECMO) initiation before ROSC, unidentifiable compression site, or poor quality/missing TEE images. Patients were divided into AV-compressed or uncompressed groups based on initial TEE findings. Documented patient characteristics, TEE recordings, resuscitation data, and critical time points were analyzed. Primary outcome was sustained ROSC. Secondary outcomes included end-tidal carbon dioxide (EtCO_2_) level at the 10th-minute post-ED arrival, any ROSC, survival to admission and discharge, active withdrawal post-resuscitation, and favorable neurological outcomes at discharge. Sample size was pre-estimated at 37 patients/group.

**Results:** From October 2020 to January 2023, 76 patients were enrolled (39 and 37 patients in the AV-uncompressed and compressed groups, respectively). Intergroup baseline characteristics were similar. The AV-uncompressed group had better probability of sustained ROSC (53.8% vs. 24.3%, odds ratio [OR] 3.63, adjusted OR [aOR] 4.72, *P*=0.010), any ROSC (56.4% vs. 32.4%, OR 2.70, aOR 3.30, *P*=0.033), and survival to admission (33.3% vs. 8.1%, OR 5.67, aOR 6.74, *P*=0.010) than the AV-compressed group. The 10th-minute EtCO_2_ levels (16.0 vs. 14.0 mmHg), active withdrawal post-resuscitation (7.7% vs. 5.4%), and survival to discharge (5.1% vs. 0%) revealed no significant intergroup differences. No patient was discharged with favorable neurological outcomes. An uncompressed AV remained an essential factor for sustained ROSC across all predefined subgroups.

**Conclusions:** Uncompressed AV during CPR increases the chances of ROSC and survival to admission among patients experiencing OHCA. However, its potential impact on long-term survival and neurological outcomes remains unclear.

**Clinical Trial Registration:** This trial was registered at ClinicalTrials.gov, identifier NCT05932784. URL: https://clinicaltrials.gov/study/NCT05932784.

**Clinical Perspective:**

1) What is new?

- When performing cardiopulmonary resuscitation according to the current guideline-recommended site, chest compressions may lead to accidental compression of the aortic valve (AV), which obstructs blood flow and worsens prognosis for patients experiencing out-of-hospital cardiac arrest.
- We have used resuscitative transesophageal echocardiography to elucidate the impact of compressed vs. uncompressed AV on outcomes of these patients, a hitherto unexplored aspect.
- Primary outcome was sustained return of spontaneous circulation (ROSC) and secondary outcomes included end-tidal carbon dioxide level at the 10th-minute post-emergency department arrival, any ROSC, survival to admission and discharge, active withdrawal post-resuscitation, and favorable neurological outcomes at discharge.
2) What are the clinical implications?

- The AV-uncompressed group had a better chance of sustained ROSC, any ROSC, and survival to admission than the AV-compressed group.
- However, its potential impact on long-term survival and neurological outcomes remains unclear; if resuscitative transesophageal echocardiography can be successfully used or if more convenient and lightweight tools can detect AV compression, both in prehospital situations, stronger evidence may be obtained.
- Current cardiopulmonary resuscitation guidelines may need to be revised for a more individualized approach, which can help rescuers avoid accidental AV compression and improve patient outcomes and prognosis.

## Introduction

Out-of-hospital cardiac arrest (OHCA) is a significant global health issue. ^1, 2^ Despite improvements in emergency medical systems and resuscitation science, the increased annual survival rate is inadequate. ^3^ More than 90% of patients experiencing OHCA did not survive to discharge, ^4^ indicating room for potential improvement in cardiac arrest management.

Chest compression is the most crucial component of post-cardiac arrest resuscitation. “Push hard, push fast, allow the chest to fully recoil, and avoid interruptions” is the optimal chest compression technique. ^5, 6^ Compression rate and depth and location are essential considerations. Current cardio-pulmonary resuscitation (CPR) guidelines recommend that chest compressions should be applied on “the center of the chest” or “the bottom half of the sternum” for all patients with cardiac arrest. ^5, 6^ However, because of inherent differences in body size or the presence of chronic heart-lung diseases, the anatomical location of the heart varies across individuals. ^7, 8^ Therefore, applying compressions at a particular site for all patients with cardiac arrest may be arbitrary. Recent observational research suggests that if chest compressions are applied at the aforementioned recommended region, more than 50% of patients experiencing OHCA may receive compressions on the left ventricular outflow tract (LVOT) or aortic root (AoR), which are located near the aortic valve (AV). ^9^

Theoretically, compressions on the LVOT or AoR during CPR could result in AV deformity, which may obstruct blood flow from the heart and severely limit coronary, cerebral, and even systemic perfusion, thereby worsening the prognosis. Animal trials have confirmed this hypothesis and demonstrated that compressions on the LVOT would significantly diminish the probability of survival. ^10^ An observational study of patients experiencing OHCA who received extracorporeal CPR (ECPR) found a 0% (0/11 cases) chance of return of electromechanical cardiac activity after extracorporeal membrane oxygenation (ECMO) flow was initiated when chest compressions were applied to the LVOT during CPR. ^11^ However, no human studies have determined whether chest compression-induced AV deformity can lead to adverse clinical outcomes and its effect on return of spontaneous circulation (ROSC), survival, or neurological outcomes to date. Resuscitative transesophageal echocardiography (TEE) provides real-time cardiac images during ongoing chest compressions and is the tool of choice for determining AV compression. ^12^

In this study, we aimed to use TEE to comprehensively analyze the effect of AV compression in patients experiencing OHCA. We hypothesized that regardless of LVOT or AoR compression, chest compressions on the AV deform it and may lead to worsening ROSC rate for these patients.

## Methods

### Data Availability Statement

The datasets generated and/or analyzed during the current study are available from the corresponding author on reasonable request.

### Study design and setting

This prospective single-center observational cohort study included all patients experiencing OHCA who underwent resuscitative TEE in the emergency department (ED) of the Far Eastern Memorial Hospital (FEMH). The FEMH is the largest tertiary medical center in New Taipei City, Taiwan, with 1415 beds and approximately 10 000 monthly ED visits, 25 of which are non-traumatic OHCAs. ^13^ The ED has been equipped with TEE for several years, and it is standard practice to perform resuscitative TEE, provided that a physician with expertise in resuscitative TEE was on duty and staffing resources permitted. In addition to being a cardiac arrest center, the FEMH offers 24-h on-call services of ECMO with formal ECPR protocol, resuscitative endovascular balloon occlusion of the aorta (REBOA), emergency coronary interventions, life-saving surgery, and target temperature management (TTM).

### Selection of participants

All patients aged ≥20 years who presented at the ED with non-traumatic OHCA and underwent resuscitative TEE were enrolled in this study. The exclusion criteria is as follows: (1) early ROSC before obtaining TEE image; (2) REBOA before ROSC (REBOA would significantly increase the hemodynamics during CPR and possibly change the likelihood of ROSC^14, 15^); (3) initiation of ECMO flow before ROSC; (4) compression site unidentified on TEE; (5) poor quality TEE image; and (6) missing TEE image.

### Ethics approval and consent to participate

This study was approved by the Institutional Review Board (IRB) of the FEMH study (IRB number: 109070-F). The Declaration of Helsinki mandates informed consent for any study involving human participants. ^16^ However, obtaining informed consent from unconscious patients is challenging when conducting cardiac arrest studies. ^17^ To facilitate vital advancements in resuscitation science, the United States Food and Drug Administration has provided specific provisions for waiving informed consent in emergency medical research. ^18^ Accordingly, the IRB approved this study including a waiver that was applicable regardless of the patients’ survival or neurological recovery. Furthermore, the observational nature of this study and the routine use of resuscitative TEE at the FEMH’s ED supported this waiver.

### Protocol of the pre- and in-hospital resuscitation

Upon identifying an emergency call as an OHCA event, the dispatcher would activate the code and notify the hospital to prepare in advance. Prior to ED arrival, as per the protocol of the New Taipei City Fire Department, at least 2 emergency medical technicians (EMTs) provided basic or advanced life support to all patients experiencing OHCA. This protocol included mechanical chest compression (LUCAS® 2, Stryker Medical, Kalamazoo, MI) at “the center of the chest” with a rate of 100 compressions/min and a depth of 5 cm, advanced airway management (endotracheal intubation or Supralaryngeal Airway), defibrillation, and administration of resuscitative medications, such as epinephrine, if the intra-venous or intra-osteo routes were available. After the initial cycle of CPR, if there was no prior “Do Not Resuscitate” order, the patient was transported to the hospital for further resuscitation.

Once the patient arrived at the ED, the rescue team assumed the resuscitation efforts. The in-hospital resuscitation was administered in accordance with the American Heart Association’s CPR guidelines. ^6^ The patient initially received chest compressions from the same machine brought by the EMTs, with the same rate and depth and at the same site. Additionally, the patient was intubated and provided mechanical ventilation with volume assist-control mode (tidal volume: 6–8 ml/kg, peak flow: 50 L/min, maximum peak pressure: 120 cm H_2_O, respiratory rate: 10/min, inspired fraction of oxygen: 100%, and positive end-expiratory pressure: 5 mmHg). After establishing an endotracheal airway, if no contraindications existed and sufficient manpower was available, TEE (PET-512MC transducer, Toshiba, Japan) was performed to identify the area of maximal compression (AMC) and to investigate potential causes of arrest. The TEE provider did not lead the code and consequently, could not interrupt resuscitation; however, the team leader was informed of the TEE findings to guide the treatment goal. If the AMC appeared positioned on the AV, the team leader decided on whether to relocate the compression site, depending on the situation. In addition, the ECMO or REBOA team were notified, if the associated criteria were met. After ROSC, the patient was transferred to the intensive care unit for post-arrest care.

### Definition of AV compression and protocol of TEE scanning

In this study, AV compression or compressed AV was defined as AV deformity caused by chest compressions detected via both long- and short-axis TEE at the mid-esophageal level using our team’s previously published algorithm. ^19^ All physicians and residents performing resuscitative TEE were required to attend a full-day workshop and clear the subsequent examination. Resuscitative TEE was applied in accordance with our team’s previously published protocol. ^20^ Two authors, J-T Sun and S-E Chu, both resuscitation and ultrasound instructors certified by Taiwan Society of Emergency Medicine and Taiwan Society of Ultrasound in Medicine, with over fifty times of experience in resuscitative TEE, reviewed all TEE images and videos of the entire CPR process for each enrolled patient post-resuscitation to determine AV compression; these results were used in this study’s analysis and served as feedback for the TEE operators.

### Study outcomes

The primary outcome was sustained ROSC, defined as ROSC >20 min. The secondary outcomes included end-tidal carbon dioxide (EtCO_2_) pressure during in-hospital resuscitation (with the level at the 10th minute following ED arrival as a representative measure), any ROSC (>1 min), survival to admission, active withdrawal of life-sustaining measures post-resuscitation (in response to confirmed unsolvable neurological prognosis), survival to discharge, and discharge with favorable neurological outcomes (defined as Modified Rankin Scale 0–2).

### Data collection and variable definitions

The clinical characteristics of the enrolled patients were collected in accordance with the current Utstein format for OHCA, ^21^ including age, gender, weight, height, body mass index, chronic disease, Charlson Comorbidity Index (CCI) ^22^, arrest location, witnessed or not, whether bystander CPR was performed, initial rhythm, prehospital management by the EMTs, in-hospital resuscitation, possible etiologies of arrest, and clinical outcomes. In addition, the critical time points of the resuscitation, such as the EMT response time (time from emergency call to EMT arrival on-scene), the total prehospital time (time from call to ED arrival), the time from ED arrival to initial TEE imaging, and the total CPR time (total prehospital time plus total in-hospital CPR time) were recorded. The EtCO_2_ data for each minute of the CPR was downloaded and recorded from the physiologic monitor. For review and analysis of the TEE findings, all TEE images and videos of each patient’s entire CPR process were captured and stored.

### Sample size determination

In Catena et al.’s study, the probability of return to electromechanical cardiac activity after the initiation of ECMO was 0% and 87.5% in the LVOT-compressed (0/11 cases) and uncompressed (7/8 cases) groups, respectively. ^11^ An animal study including 26 swine revealed that the rate of ROSC was 0% (0/13 cases) and 69.2% (9/13 cases) in the LVOT-compressed and the left-ventricle (LV) compressed groups, respectively. ^10^ Nonetheless, an ROSC rate of 87.5% or 69.2% is not achievable in actual clinical practice, and the outcomes of LV compression cannot fully represent the AV-uncompressed group. Therefore, in this study, for the AV-uncompressed group, the baseline sustained-ROSC rate (25.6%) of patients experiencing OHCA in Taipei was used. ^3^ For the AV-compressed group, the sustained ROSC rate was hypothesized to be proximate to 0%. In addition, previous studies demonstrated that approximately 50% of patients undergoing CPR would experience AV compression. ^7–9^ Therefore, the enrollment ratio between these 2 groups was postulated to be 1:1. Finally, 34 patients/group were estimated to provide the trial with 90% power to detect the difference in sustained ROSC rate using an uncorrected 2-sided chi-square test at a significance level of 0.05. To compensate for missing values or unexpectable events, the number of patients was increased by 10%, to a total of 37 in each arm. The outcomes of patients whose AV was compressed during CPR and those whose AV was uncompressed were compared.

### Data analysis

In this study, the Shapiro–Wilk test was used to assess the existence of a normal distribution for all continuous data. Data with a normal distribution were recorded as the mean with standard deviation and analyzed with 2-tailed T-tests. Data without a normal distribution were presented as the median with interquartile range and analyzed with a 2-sided Mann–Whitney U test. Numeric variables, such as CCI and epinephrine administration times, were presented as median with interquartile range and examined using the 2-sided Mann–Whitney U test. Categorical variables were expressed as frequencies with corresponding percentages and assessed using chi-square tests. The odds ratio (OR) and 95% CIs of the study’s outcomes between the 2 groups were calculated using univariate logistic regression and adjusted for the associated confounders using multivariate logistic regression. The confounders included variables that have been confirmed to have a significant association with our study outcomes, as reported in previous studies, such as sex, ^23, 24^ CCI, ^25–27^ witnessed arrest, ^4^ bystander CPR, ^4^ initial shockable rhythm, ^28, 29^ total prehospital time. ^30, 31^ Post-hoc subgroup analyses were conducted using multivariate logistic regression in pre-defined groups, including initial shockable rhythm, witnessed collapse, bystander CPR, cardiogenic arrest, patients’ CCI, age, total prehospital time, and EMT response time. All subgroup models were adjusted for sex, CCI, witnessed arrest, bystander CPR, initial shockable rhythm, and total prehospital time, except for the variables themselves. Some variables had zero cell counts; therefore, the first regression method was utilized in the model. The model’s goodness-of-fit was assessed using the Hosmer–Lemeshow test. To further analyze the EtCO_2_ differences between the 2 groups, additional generalized estimating equations (GEE) models were employed to examine the differences across cumulative time periods. In this study, the limited missing data were addressed by eliminating rows or columns with absent values, thereby preserving the overall integrity of our dataset. All statistical analyses were conducted using the IBM® SPSS® Statistics version 26.0. A 2-tailed *P*-value of <0.05 was determined to be statistically significant.

### Independent Data Access and Analysis

This study was conducted at Far Eastern Memorial Hospital, with data collection handled onsite. The task of data access and analysis was entrusted to the independent Statistical Consulting Unit at National Taiwan University Hospital. Importantly, these institutions operate separately, with no organizational affiliation, ensuring the absence of conflicts of interest. Data analysis was conducted in a strictly independent manner, with analysts blind to the personnel at Far Eastern Memorial Hospital. This autonomy in data processing fortifies the objectivity of our results, enhancing both data validity and participant safety in the trial.

## Results

From October 2020 to January 2023, 104 patients experiencing OHCA underwent ED resuscitative TEE Twenty-eight patients did not meet the inclusion criteria and were excluded; 8 patients achieved ROSC prior to obtaining TEE images, 1 was diagnosed with a ruptured aortic aneurysm via bedside ultrasound and underwent REBOA, 4 met the ECPR criteria and were resuscitated with ECMO before ROSC, 3 had type-A aortic dissection-induced cardiac tamponade diagnosed via TEE making it difficult to identify the AMC, 4 had poor TEE images, and 8 TEE images were missing, hindering the program directors’ review of the videos to confirm the AMC. The trial was temporarily suspended in May and June of 2020 and in May and June of 2021 because of the coronavirus disease 2019 pandemic. The final trial cohort comprised 39 and 37 patients in the AV-uncompressed and compressed groups, respectively **(Figure 1)**. Furthermore, during the pre-analysis stage, missing prehospital time (due to an EMT recording error) for 1 patient and inconsistencies in the EtCO_2_ data for 3 other patients were identified. These four instances of data were accordingly handled as missing data.

**Figure 1.**
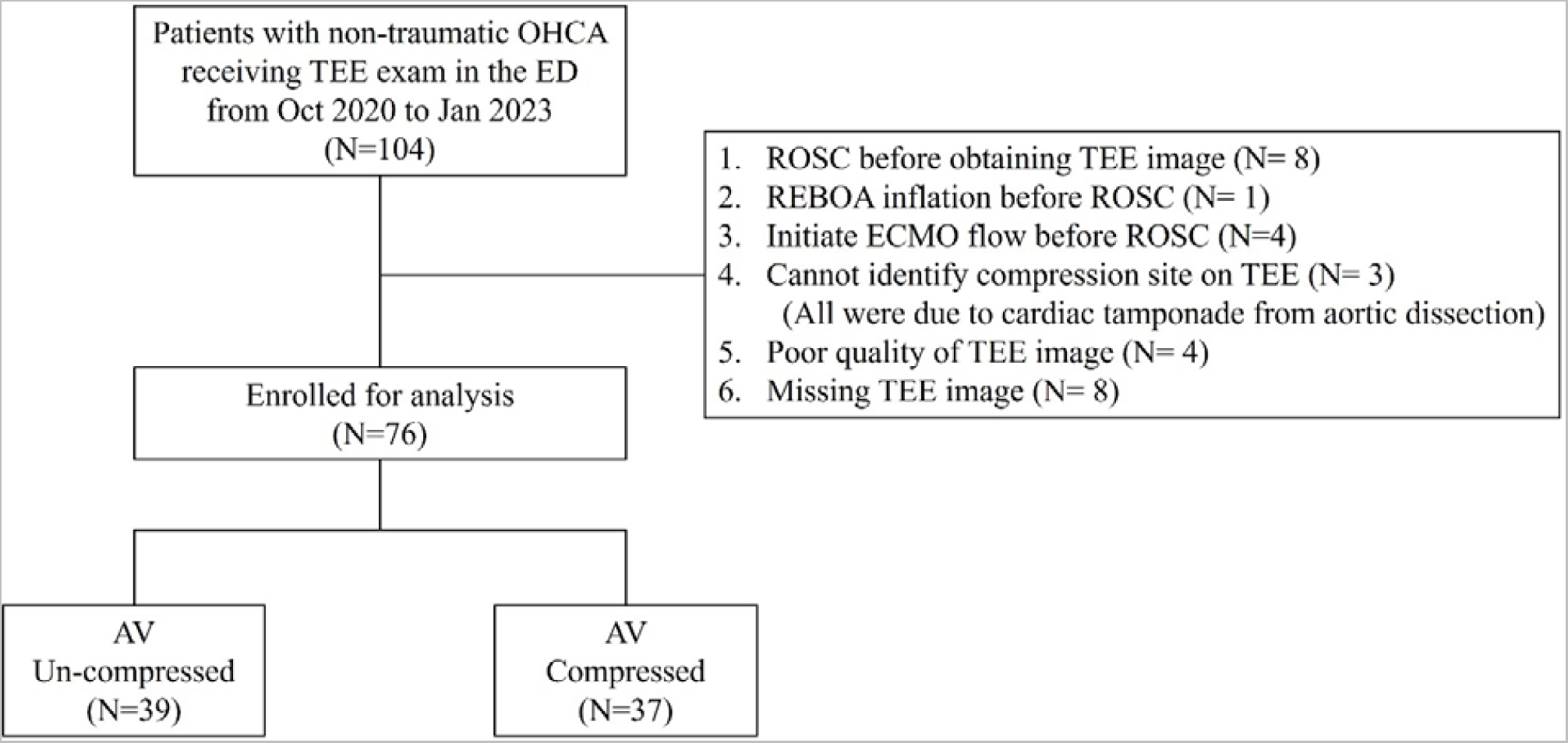
Flowchart of the study OHCA, Out-of-hospital cardiac arrest; TEE, transesophageal echocardiography; ED, emergency department; ROSC, return of spontaneous circulation; ECMO, extracorporeal membrane oxygenation; REBOA, resuscitative endovascular balloon occlusion of the aorta; AV, aortic valve.

The enrolled patients had a mean age of 65 years, and most were males (67.1%). Approximately 17.1% of patients experienced cardiac arrest in public settings and 31.6% of these arrests were witnessed. Nearly 50% of patients received bystander’s CPR. Approximately, 19.7% of patients initially presented with a shockable rhythm and were defibrillated on-scene. Before undergoing TEE, both groups of patients had already received approximately 30 min of CPR. No statistically significant intergroup differences were observed in terms of patient demographics, body mass index, underlying comorbidities, locations or causes of cardiac arrest, or pre- and in-hospital treatments **(Table 1)**.

**Table 1.**
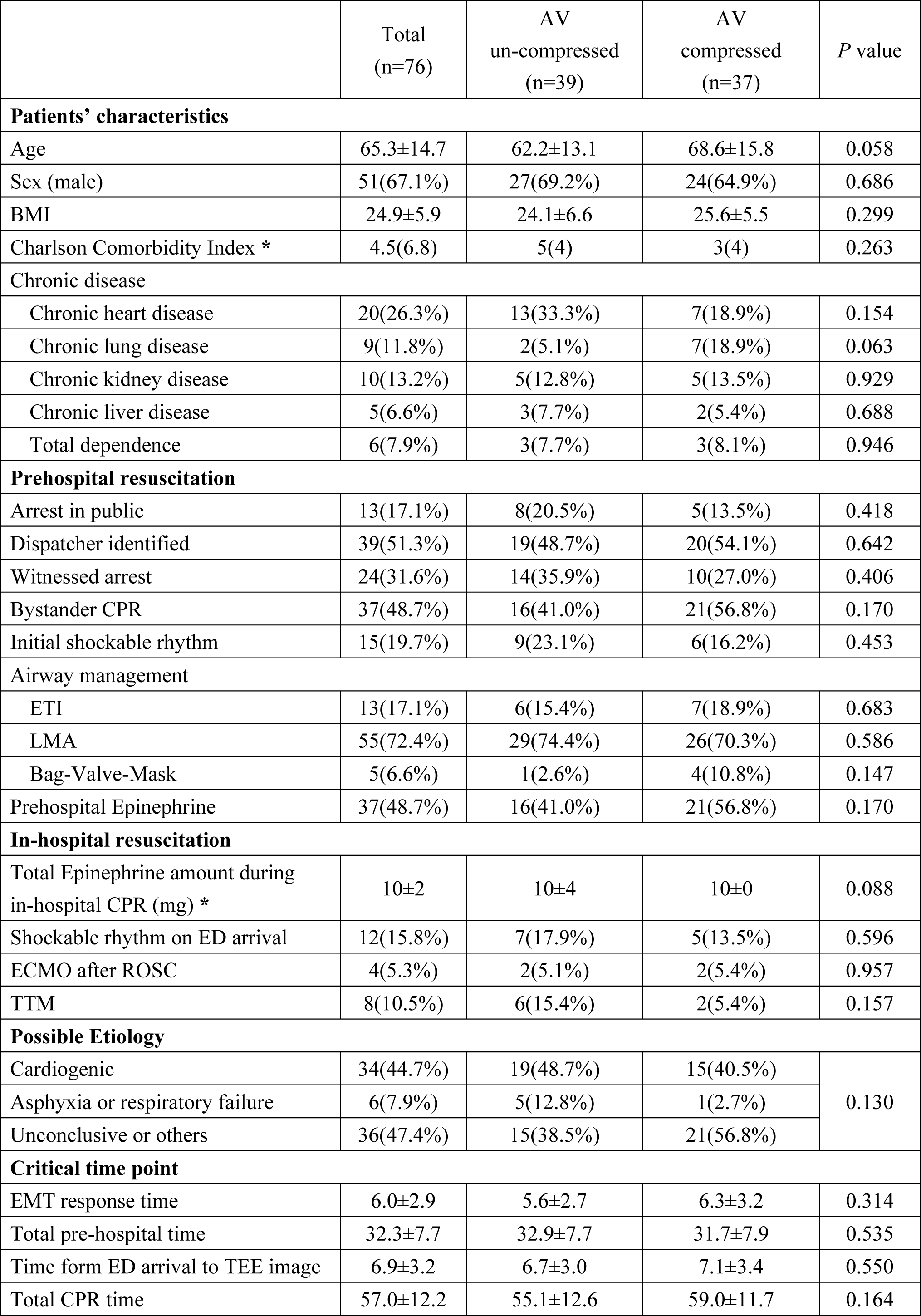
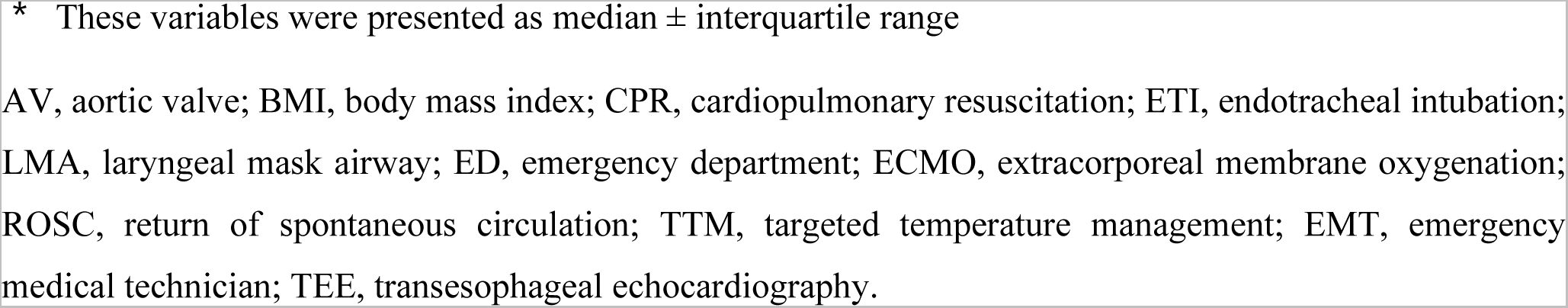
Baseline characteristics of the study participants.

|  | Total<br>(n=76) | AV<br>un-compressed<br>(n=39) | AV<br>compressed<br>(n=37) | *P* value |
| --- | --- | --- | --- | --- |
| **Patients' characteristics** |  |  |  |  |
| Age | 65.3±14.7 | 62.2±13.1 | 68.6±15.8 | 0.058 |
| Sex (male) | 51(67.1%) | 27(69.2%) | 24(64.9%) | 0.686 |
| BMI | 24.9±5.9 | 24.1±6.6 | 25.6±5.5 | 0.299 |
| Charlson Comorbidity Index * | 4.5(6.8) | 5(4) | 3(4) | 0.263 |
| Chronic disease |  |  |  |  |
| Chronic heart disease | 20(26.3%) | 13(33.3%) | 7(18.9%) | 0.154 |
| Chronic lung disease | 9(11.8%) | 2(5.1%) | 7(18.9%) | 0.063 |
| Chronic kidney disease | 10(13.2%) | 5(12.8%) | 5(13.5%) | 0.929 |
| Chronic liver disease | 5(6.6%) | 3(7.7%) | 2(5.4%) | 0.688 |
| Total dependence | 6(7.9%) | 3(7.7%) | 3(8.1%) | 0.946 |
| **Prehospital resuscitation** |  |  |  |  |
| Arrest in public | 13(17.1%) | 8(20.5%) | 5(13.5%) | 0.418 |
| Dispatcher identified | 39(51.3%) | 19(48.7%) | 20(54.1%) | 0.642 |
| Witnessed arrest | 24(31.6%) | 14(35.9%) | 10(27.0%) | 0.406 |
| Bystander CPR | 37(48.7%) | 16(41.0%) | 21(56.8%) | 0.170 |
| Initial shockable rhythm | 15(19.7%) | 9(23.1%) | 6(16.2%) | 0.453 |
| Airway management |  |  |  |  |
| ETI | 13(17.1%) | 6(15.4%) | 7(18.9%) | 0.683 |
| LMA | 55(72.4%) | 29(74.4%) | 26(70.3%) | 0.586 |
| Bag-Valve-Mask | 5(6.6%) | 1(2.6%) | 4(10.8%) | 0.147 |
| Prehospital Epinephrine | 37(48.7%) | 16(41.0%) | 21(56.8%) | 0.170 |
| **In-hospital resuscitation** |  |  |  |  |
| Total Epinephrine amount during in-hospital CPR (mg) * | 10±2 | 10±4 | 10±0 | 0.088 |
| Shockable rhythm on ED arrival | 12(15.8%) | 7(17.9%) | 5(13.5%) | 0.596 |
| ECMO after ROSC | 4(5.3%) | 2(5.1%) | 2(5.4%) | 0.957 |
| TTM | 8(10.5%) | 6(15.4%) | 2(5.4%) | 0.157 |
| **Possible Etiology** |  |  |  |  |
| Cardiogenic | 34(44.7%) | 19(48.7%) | 15(40.5%) | 0.130 |
| Asphyxia or respiratory failure | 6(7.9%) | 5(12.8%) | 1(2.7%) |  |
| Unconclusive or others | 36(47.4%) | 15(38.5%) | 21(56.8%) |  |
| **Critical time point** |  |  |  |  |
| EMT response time | 6.0±2.9 | 5.6±2.7 | 6.3±3.2 | 0.314 |
| Total pre-hospital time | 32.3±7.7 | 32.9±7.7 | 31.7±7.9 | 0.535 |
| Time form ED arrival to TEE image | 6.9±3.2 | 6.7±3.0 | 7.1±3.4 | 0.550 |
| Total CPR time | 57.0±12.2 | 55.1±12.6 | 59.0±11.7 | 0.164 |

\* These variables were presented as median ± interquartile range
AV, aortic valve; BMI, body mass index; CPR, cardiopulmonary resuscitation; ETI, endotracheal intubation;
LMA, laryngeal mask airway; ED, emergency department; ECMO, extracorporeal membrane oxygenation;
ROSC, return of spontaneous circulation; TTM, targeted temperature management; EMT, emergency
medical technician; TEE, transesophageal echocardiography.

Among the 76 enrolled patients, 34 (44.7%) achieved ROSC, with 30 (39.5%) maintaining sustained ROSC. Additionally, 16 (21.1%) patients survived until hospital admission, 5 (6.6%) had life-sustaining measures withdrawn during hospitalization for their irreversible neurological prognosis, and 2 (2.6%) survived until discharge. However, none exhibited a favorable neurological recovery, irrespective of AV compression. The AV-uncompressed group had a significantly higher sustained ROSC rate (53.8% vs. 24.3%, unadjusted OR 3.63, *P*=0.010), as well as in any ROSC (*P*=0.038), and survival to admission (*P*=0.012) than the AV-compressed group **(Table 2)**. The proportion of active withdrawal after resuscitation was similar between both groups (unadjusted OR 1.46, *P*=0.689). Compared with 2 patients in the AV-uncompressed group, no patient survived to discharge in the AV-compressed group; however, this difference was not statistically significant. For the potential confounders associated with study’s outcomes, additional multivariate logistic regression analyses were performed **(Table 2)**. An uncompressed AV remains a statistically significant factor in achieving sustained ROSC (*P*=0.009), including in any ROSC (*P*=0.033), and survival to hospital admission (*P*=0.010). **Figure 2** presents the subgroup analysis, revealing that in all predefined subgroups, an uncompressed AV is an important factor for sustained ROSC.

**Figure 2.**
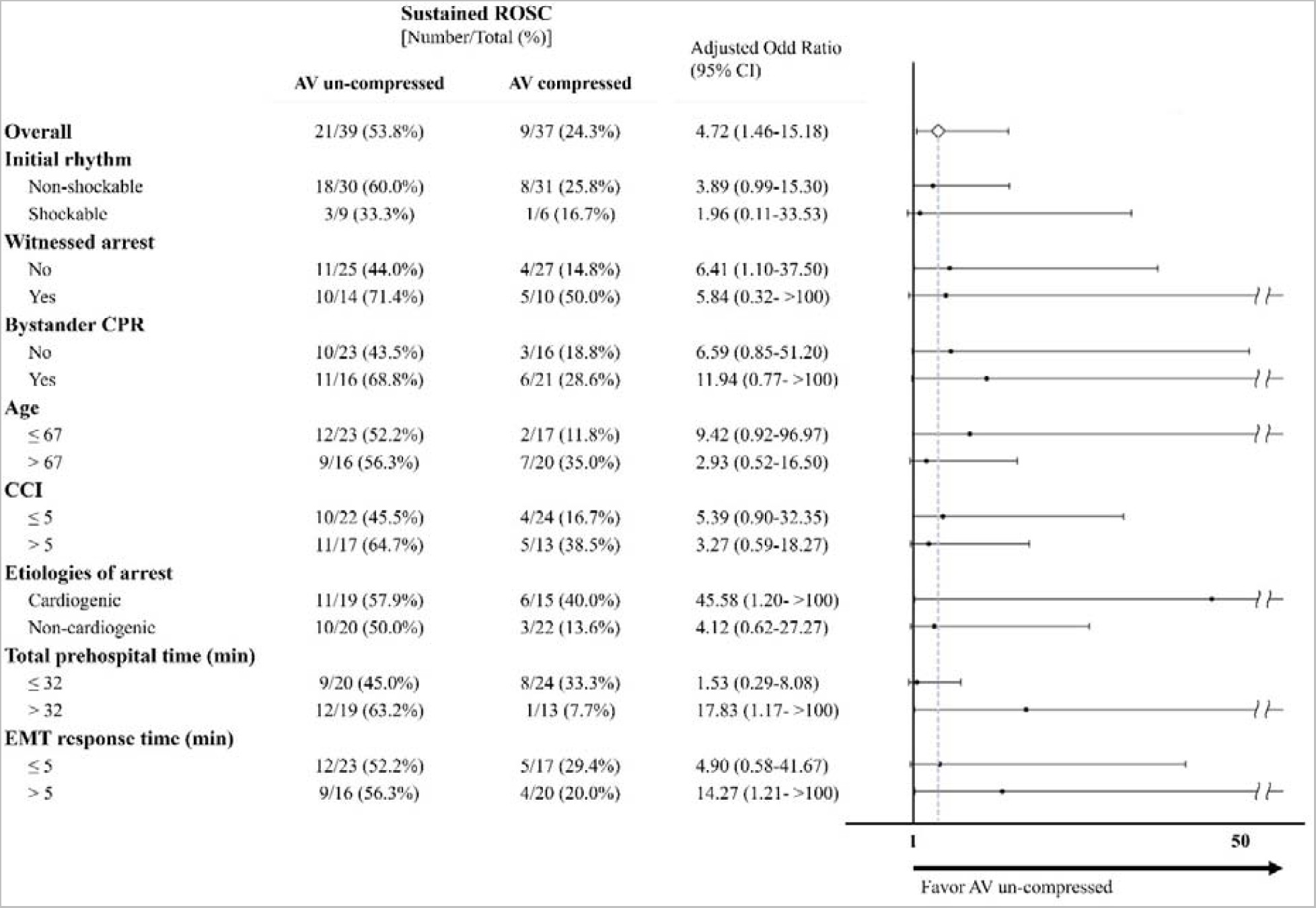
Subgroup results for sustained ROSC In the subgroup analysis model for age, CCI, total pre-hospital time, and EMT response time, the cut-off values are determined based on the medians of the enrolled population. All models are adjusted for sex, CCI, witnessed arrest, bystander CPR, initial shockable rhythm, and total prehospital time, except for the variables themselves. The blue dashed line represents the overall OR value. ROSC, return of spontaneous circulation; AV, aortic valve; CPR, cardiopulmonary resuscitation; CCI, Charlson Comorbidity Index; EMT, emergency medical technician; OR, odds ratio.

**Table 2.** Primary and Secondary outcomes.

|  | AV<br>un-compressed<br>(n=39) | AV<br>compressed<br>(n=37) | Unadjusted<br>Odds Ratio<br>(95% CI) | *P* value | Adjusted<br>Odds Ratio<br>(95% CI) | *P* value |
| --- | --- | --- | --- | --- | --- | --- |
| **Primary Outcomes** |  |  |  |  |  |  |
| Sustained ROSC | 21 (53.8%) | 9 (24.3%) | 3.63<br>(1.36-9.67) | **0.010** | 4.72<br>(1.46-15.18) | **0.009** |
| **Secondary Outcomes** |  |  |  |  |  |  |
| EtCO<sub>2</sub> level at the 10th<br>minute post-ED arrival | *16.0<br>(10.0-25.0) | *14.0<br>(3.75-36.75) | 0.99<br>(0.97-1.02) | 0.673 | 0.99<br>(0.97-1.02) | 0.674 |
| Any ROSC | 22 (56.4%) | 12 (32.4%) | 2.70<br>(1.06-6.87) | **0.038** | 3.30<br>(1.10-9.88) | **0.033** |
| Survive to<br>hospital admission | 13 (33.3%) | 3 (8.1%) | 5.67<br>(1.46-21.97) | **0.012** | 6.74<br>(1.59-28.50) | **0.010** |
| Active withdrawal<br>after resuscitation | 3 (7.7%) | 2 (5.4%) | 1.46<br>(0.23-9.26) | 0.689 | 1.38<br>(0.21-9.10) | 0.741 |
| Survive to<br>hospital discharge | 2 (5.1%) | 0 (0.0%) | N/A | N/A | N/A | N/A |
| Discharge with<br>mRS ≤ 2 | 0 (0.0%) | 0 (0.0%) | N/A | N/A | N/A | N/A |

\* These cells were presented as median with first and third quartiles (Q1– Q3).
All models are adjusted for sex, Charlson Comorbidity Index, witnessed arrest, bystander CPR, initial shockable rhythm, and total pre-hospital time.
ROSC, return of spontaneous circulation; EtCO<sub>2</sub>, end-tidal carbon dioxide; ED, emergency department; AV, aortic valve; mRS, modified Rankin scale; N/A, not applicable.

In the assessment of EtCO_2_, no statistically significant divergence was observed in the levels recorded at the 10th minute following ED arrival between both groups **(Table 2)**. Additionally, throughout the in-hospital resuscitation phase, despite notable differences in the patterns of EtCO_2_ levels between both groups **(Figure 3)**, where the AV-uncompressed group tended to display higher levels in the initial 10 min, and the AV-compressed group displayed a higher tendency between the 15th and 25th min, no statistically significant differences were found in the minute-to-minute EtCO2 levels **(Table S1)**. A subsequent post-hoc GEE model analysis indicated that no marked difference in EtCO_2_ levels between both groups emerged within the first 10 min, irrespective of the chosen cut-off point **(Table S2)**.

**Figure 3.**
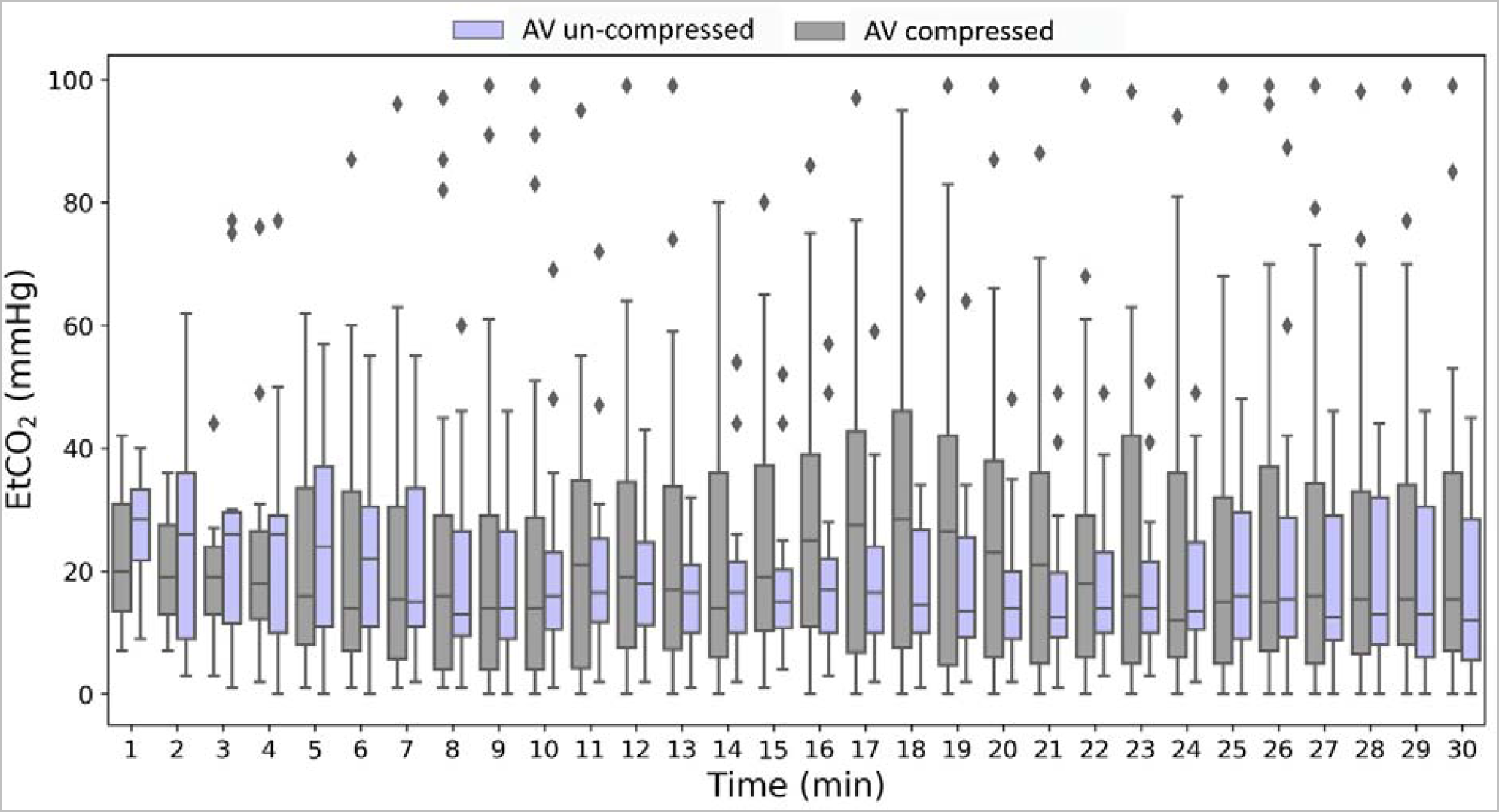
EtCO_2_ levels at each minute of resuscitation after ED arrival All data points are presented as median and interquartile range. EtCO_2_, end-tidal carbon dioxide; ED, emergency department.

## Discussion

In our study, we discovered that the chest compression site is essential in determining the prognosis of patients experiencing OHCA. Our study results indicated that applying chest compressions directly on the AV considerably reduced the chances of achieving ROSC, sustained ROSC, and survival to admission. Thus, our study has underscored the importance of avoiding direct AV compression during CPR for patients experiencing OHCA for an improved prognosis.

To the best of our knowledge, this is the first prospective study to investigate the relationship between chest compression site and outcomes such as ROSC and survival in patients experiencing OHCA. Our findings might be attributed to the potential adverse impact of chest compressions on AV movement that reduces blood flow from the LV to the AoR. This hypothesis is credible since it directly influences the perfusion of the coronary arteries, which originate from the AoR and perfuse the myocardium. In Anderson et al.’s animal study, the AV-compressed group had considerably lower systolic and diastolic aortic pressures during resuscitation, and a significant drop in coronary perfusion pressure during the advanced life support phase. ^10^ More in-depth human studies on the CPR hemodynamics may be necessary in the future to further establish the connection between compression site and patient prognosis.

This study’s findings revealed that the ratio of compressed to uncompressed AV is proximate to 1:1, consistent with previously reported findings. ^7–9, 32^ A recent observational study reported that if chest compressions were performed in the guideline-recommended position, 53% of patients experiencing OHCA may have AV compression (9/17 cases). ^9^ Hwang et al. reported an incidence of 79% of AV compression in patients experiencing OHCA (27/34 cases). ^32^ These evidences suggest that when performing CPR according to current guidelines, majority of the resuscitation attempts may be ineffective or potentially harmful because of direct compression of the patient’s AV. These guidelines may need to be revised for a more individualized approach, which may help rescuers to avoid accidental AV compression.

To identify the optimal compression site during CPR, several imaging studies demonstrated that the ventricles and the AV were mainly positioned beneath “the lower third of the sternum,” and “the center of the chest,” respectively. ^7, 8^ Subsequently, Cha et al.’s study to evaluate the effects of different compression sites revealed that compressions at the “lower end of the sternum” resulted in higher EtCO_2_ than compressions at the standard “center of the chest.” ^33^ However, Qvigstad et al.’s study did not reach a similar conclusion. No significant difference in EtCO_2_ was observed regardless of whether the compression site was at the midpoint of the inter-nipple line (INL), 2 cm below the INL, 2 cm below and to the left of the INL, or 2 mc below and to the right of the INL. ^34^ Therefore, the usefulness of revising the standard compression site to the “lower third of sternum” remains uncertain. The significant differences between the aforementioned studies have 2 possible explanations.

First, EtCO_2_ level may not be a suitable hemodynamic indicator for OHCA research. Among patients experiencing OHCA, EtCO_2_ levels might vary widely, which could be due to the quality of CPR and the etiologies or duration of cardiac arrest, ^35, 36^ varying lung volumes and pulmonary compliance, ^37, 38^ or even measuring artifacts on the physiological monitor. ^39, 40^ In our study, similar to the findings of Qvigstad et al., no significant intergroup difference in EtCO_2_ was observed throughout the in-hospital CPR process. This lack of distinction could be attributed to the aforementioned confounding variables and the fact that both groups had already undergone approximately 30 min of resuscitation prior to ED arrival. Insufficient sample size, which may have prevented the generation of statistically significant differences, is another possible factor. Moreover, the varying distribution of patients achieving ROSC over time within these groups could contribute to the challenges in using EtCO_2_ as a reliable indicator of CPR quality **(Figure S1)**. Thus, EtCO_2_ might serve as a supplementary assessment rather than as a primary indicator when determining the precise effectiveness of various compression sites in OHCA research. Therefore, in our study, we used a more stable and reliable indicator, “sustained ROSC,” as a primary outcome and included an adequate sample size to examine the intergroup differences. Further research is required to evaluate the effect of various compression sites on survival and neurological outcomes.

Second, the effectiveness of preventing AV compression cannot be guaranteed by mere reliance on body landmarks. Variations in individual body shape and medical history could result in differences in anatomical cardiac positions, leading to diverse hemodynamic responses during CPR, despite chest compressions being applied at similar body landmarks. Particularly, in Qvigstad et al.’s study, the difference in chest compression sites were merely 2 cm apart. ^34^ This minor discrepancy in distance may confound the generation of a meaningful difference in hemodynamics, let alone completely avoid AV compression. In this study, TEE was used to accurately determine the occurrence of AV compression during CPR. In the future, TEE augmented by improved technologies may help rescuers precisely locate the AV to ensure more efficient chest compressions.

Within the AV-compressed group in this study, some rescuers adjusted the chest compression site based on patient conditions or available resources in an attempt to decompress the AV. This led to crossover situations; however, as this was observational research that did not interfere with clinical decisions, and in line with the “intention to treat” principle, patients whose chest compression site was adjusted were not excluded. Therefore, an additional analysis was conducted on the AV-compressed group to account for this deviation **(Table S3)**. Out of 37 patients, the AV became uncompressed in 14 patients following TEE-guided compression site adjustment, whereas in 23 patients, the AV remained persistently compressed regardless of any adjustment attempts. Notably, regardless of whether the AV was ultimately uncompressed, we did not observe a noticeable difference in outcomes. Furthermore, the prognosis for the AV-compressed group remained significantly worse compared with that for the AV-uncompressed group, despite these adjustments in compression site. This might be attributed to our cohort undergoing prolonged CPR; on an average, it involved approximately 30 min of prehospital resuscitation and an additional 11 min post-ED arrival resuscitation to successfully uncompress the AV. Under such extended resuscitation scenarios, any intervention might be less effective, thus explaining the observed results. Further research should evaluate the clinical impact of early TEE-guided compression site adjustment, even in prehospital settings or for patients with in-hospital cardiac arrest.

This study, primarily focused on patients experiencing non-traumatic OHCA, has underscored the detrimental effects of AV compression on ROSC and overall survival outcomes. However, generalization of these findings to traumatic OHCA cases is not recommended. Notably, a study conducted on a traumatic OHCA swine model revealed that chest compressions near the AV (or LVOT) during resuscitation significantly diminished coronary perfusion and arterial pressures and EtCO_2_ levels, thereby drastically reducing the survival rate. ^41^ Despite these findings, clinical data affirming the adverse effects of AV compression in patients with traumatic cardiac arrest is lacking, highlighting the need for further research in this domain. In addition, the potential for similar effects in patients experiencing in-hospital cardiac arrest and the potential benefits of prehospital TEE-guided chest compressions remain largely unexplored. Therefore, more extensive research efforts must focus on these areas in the future.

### Limitations

This study has some limitations. First, performing TEE outside the hospital remains a challenge; hence, it is unclear if the initial TEE findings, whether AV was being compressed, were consistent with those during prehospital resuscitation. However, since we did not change the compression device, position, or depth post-ED arrival, it is probable that the AMC observed initially on TEE at the ED approximates the AMC during prehospital resuscitation. In the future, if TEE can be successfully used or if more convenient and lightweight tools can detect AV compression, both in prehospital situations, stronger evidence may be obtained. Second, as our study design excluded patients who achieved early ROSC prior to TEE and those who were resuscitated with ECMO or REBOA, our cohort primarily comprised patients who had already undergone prolonged CPR. This specific selection criteria might have inadvertently led to an underrepresentation of long-term outcomes, such as survival rates until hospital discharge and the associated neurological outcomes at discharge. A more extensive study incorporating prehospital TEE might potentially to address this limitation. Third, constrained by staffing resources, approximately only 23% of patients experiencing OHCA arriving at the ED during the study period were able to undergo resuscitative TEE. Despite concerns of potential selection bias, this rate of resuscitative TEE aligns with those previously reported, ^9, 42^ indicating a common challenge in clinical settings. Nevertheless, this limitation highlights the need for further extensive studies to minimize selection bias and provide a more comprehensive understanding of the issue.

## Conclusion

In patients experiencing OHCA, CPR performed without compressing the AV considerably increases the probability of achieving ROSC, sustaining ROSC, and survival to hospital admission. However, the beneficial impact of an uncompressed AV on long-term survival and neurological outcomes requires further exploration and research.

## Acknowledgements

We thank the nursing team from the Departments of Emergency Medicine Critical Care at Far Eastern Memorial Hospital for their outstanding resuscitation efforts and post-ROSC care given to patients experiencing OHCA. Furthermore, we express our gratitude to the staff at the National Taiwan University Hospital Statistical Consulting Unit, whose efforts considerably advanced our work.

## Disclosures

### Statistical Analysis

Formal analysis was approved by the staff, Dr. Chin-Ho Chang, of the National Taiwan University Hospital Statistical Consulting Unit.

## Supplemental Material

Tables S1–S3 Figure S1

## Non-standard Abbreviations and Acronyms

AMC: Area of Maximal Compression
AoR: Aortic root
AV: Aortic valve
CCI: Charlson Comorbidity Index
ECPR: Extracorporeal cardiopulmonary resuscitation
EtCO_2_: End-tidal Carbon Dioxide
FEMH: Far Eastern Memorial Hospital
GEE: Generalized estimating equations
INL: Inter-nipple line
mRS: Modified Rankin Scale
N/A: Not Applicable.
OHCA: Out-of-Hospital Cardiac Arrest
REBOA: Resuscitative Endovascular Balloon Occlusion of the Aorta
ROSC: Return of Spontaneous Circulation
TEE: Transesophageal Echocardiography
TTM: Target Temperature Management

## References

1. Ong ME, Shin SD, De Souza NN, Tanaka H, Nishiuchi T, Song KJ, Ko PC, Leong BS, Khunkhlai N, Naroo GY, et al. Outcomes for out-of-hospital cardiac arrests across 7 countries in Asia: the Pan Asian Resuscitation Outcomes Study (PAROS). Resuscitation. 2015;96:100–108. doi: 10.1016/j.resuscitation.2015.07.026

2. Aufderheide TP, Nolan JP, Jacobs IG, van Belle G, Bobrow BJ, Marshall J, Finn J, Becker LB, Bottiger B, Cameron P, et al. Global health and emergency care: a resuscitation research agenda—Part 1. Academic Emergency Medicine. 2013;20:1289–1296. doi: https://doi.org/10.1111/acem.12270

3. Lin HY, Chien YC, Lee BC, Wu YL, Liu YP, Wang TL, Ko PCI, Chong KM, Wang HC, Huang EPC, et al. Outcomes of out-of-hospital cardiac arrests after a decade of system-wide initiatives optimising community chain of survival in Taipei city. Resuscitation. 2022;172:149–158. doi: https://doi.org/10.1016/j.resuscitation.2021.12.027

4. Yan S, Gan Y, Jiang N, Wang R, Chen Y, Luo Z, Zong Q, Chen S, Lv C. The global survival rate among adult out-of-hospital cardiac arrest patients who received cardiopulmonary resuscitation: a systematic review and meta-analysis. Critical Care. 2020;24:61. doi: 10.1186/s13054-020-2773-2

5. Soar J, Böttiger BW, Carli P, Couper K, Deakin CD, Djärv T, Lott C, Olasveengen T, Paal P, Pellis T, et al. European Resuscitation Council Guidelines 2021: adult advanced life support. Resuscitation. 2021;161:115–151. doi: 10.1016/j.resuscitation.2021.02.010

6. Panchal AR, Bartos JA, Cabañas JG, Donnino MW, Drennan IR, Hirsch KG, Kudenchuk PJ, Kurz MC, Lavonas EJ, Morley PT, et al. Part 3: Adult Basic and Advanced Life Support: 2020 American Heart Association Guidelines for Cardiopulmonary Resuscitation and Emergency Cardiovascular Care. Circulation. 2020;142:S366–S468. doi: doi:10.1161/CIR.0000000000000916

7. Nestaas S, Stensæth KH, Rosseland V, Kramer-Johansen J. Radiological assessment of chest compression point and achievable compression depth in cardiac patients. Scand J Trauma Resusc Emerg Med. 2016;24:54. doi: 10.1186/s13049-016-0245-0

8. Shin J, Rhee JE, Kim K. Is the inter-nipple line the correct hand position for effective chest compression in adult cardiopulmonary resuscitation? Resuscitation. 2007;75:305–310. doi: 10.1016/j.resuscitation.2007.05.003

9. Teran F, Dean AJ, Centeno C, Panebianco NL, Zeidan AJ, Chan W, Abella BS. Evaluation of out-of-hospital cardiac arrest using transesophageal echocardiography in the emergency department. Resuscitation. 2019;137:140–147. doi: 10.1016/j.resuscitation.2019.02.013

10. Anderson KL, Castaneda MG, Boudreau SM, Sharon DJ, Bebarta VS. Left ventricular compressions improve hemodynamics in a swine model of out-of-hospital cardiac arrest. Prehosp Emerg Care. 2017;21:272–280. doi: 10.1080/10903127.2016.1241328

11. Catena E, Ottolina D, Fossali T, Rech R, Borghi B, Perotti A, Ballone E, Bergomi P, Corona A, Castelli A, et al. Association between left ventricular outflow tract opening and successful resuscitation after cardiac arrest. Resuscitation. 2019;138:8–14. doi: 10.1016/j.resuscitation.2019.02.027

12. Teran F, Prats MI, Nelson BP, Kessler R, Blaivas M, Peberdy MA, Shillcutt SK, Arntfield RT, Bahner D. Focused transesophageal echocardiography during cardiac arrest resuscitation: JACC Review Topic of the Week. J Am Coll Cardiol. 2020;76:745–754. doi: 10.1016/j.jacc.2020.05.074

13. Emergency Medical Services Resuscitation Success Statistics, as provided by the Fire Department of the Ministry of the Interior, Taiwan. https://www.nfa.gov.tw/cht/index.php?act=download&ids=14489. 2021.

14. Jang D-H, Lee DK, Jo YH, Park SM, Oh YT, Im CW. Resuscitative endovascular occlusion of the aorta (REBOA) as a mechanical method for increasing the coronary perfusion pressure in non-traumatic out-of-hospital cardiac arrest patients. Resuscitation. 2022;179:277–284. doi: https://doi.org/10.1016/j.resuscitation.2022.07.020

15. Gamberini L, Coniglio C, Lupi C, Tartaglione M, Mazzoli CA, Baldazzi M, Cecchi A, Ferri E, Chiarini V, Semeraro F, et al. Resuscitative endovascular occlusion of the aorta (REBOA) for refractory out of hospital cardiac arrest. An Utstein-based case series. Resuscitation. 2021;165:161–169. doi: 10.1016/j.resuscitation.2021.05.019

16. Association WM. World Medical Association Declaration of Helsinki: Ethical principles for medical research involving human subjects. JAMA. 2013;310:2191–2194. doi: 10.1001/jama.2013.281053

17. Foëx BA. The problem of informed consent in emergency medicine research. Emergency Medicine Journal. 2001;18:198–204. doi: 10.1136/emj.18.3.198

18. Biros MH, Fish SS, Lewis RJ. Implementing the Food and Drug Administration’s final rule for waiver of informed consent in certain emergency research circumstances. Acad Emerg Med. 1999;6:1272–1282. doi: 10.1111/j.1553-2712.1999.tb00144.x

19. Chang CJ, Sun JT, Ma MHM, Chiang WC, Chu SE. Precise identification of area of maximal compression using transesophageal echocardiography during cardiopulmonary resuscitation. Resuscitation. 2023;187. doi: 10.1016/j.resuscitation.2023.109804

20. Chu SE, Chang CJ, Chen HA, Chiu YC, Huang CY, Huang EPC, Hsieh MJ, Chiang WC, Ma MHM, Sun JT. Core Ultrasound in REsuscitation (CURE): a novel protocol for ultrasound-assistant life support via application of both transesophageal and transthoracic ultrasound. Resuscitation. 2022;173:1–3.

21. Perkins GD, Jacobs IG, Nadkarni VM, Berg RA, Bhanji F, Biarent D, Bossaert LL, Brett SJ, Chamberlain D, Caen ARd, et al. Cardiac arrest and cardiopulmonary resuscitation outcome reports: update of the Utstein Resuscitation Registry templates for out-of-hospital cardiac arrest. Circulation. 2015;132:1286–1300. doi: doi:10.1161/CIR.0000000000000144

22. Charlson ME, Pompei P, Ales KL, MacKenzie CR. A new method of classifying prognostic comorbidity in longitudinal studies: development and validation. J Chronic Dis. 1987;40:373–383. doi: 10.1016/0021-9681(87)90171-8

23. Lei H, Hu J, Liu L, Xu D. Sex differences in survival after out-of-hospital cardiac arrest: a meta-analysis. *Critical care (London*, England). 2020;24:613–613. doi: 10.1186/s13054-020-03331-5

24. Morrison LJ, Schmicker RH, Weisfeldt ML, Bigham BL, Berg RA, Topjian AA, Abramson BL, Atkins DL, Egan D, Sopko G. Effect of gender on outcome of out of hospital cardiac arrest in the Resuscitation Outcomes Consortium. Resuscitation. 2016;100:76–81.

25. Dumas F, Paoli A, Paul M, Savary G, Jaubert P, Chocron R, Varenne O, Mira JP, Charpentier J, Bougouin W, et al. Association between previous health condition and outcome after cardiac arrest. Resuscitation. 2021;167:267–273. doi: 10.1016/j.resuscitation.2021.06.017

26. Oving I, van Dongen LHPI, Deurholt SC, Ramdani A, Beesems SG, Tan HL, Blom MT. Comorbidity and survival in the pre-hospital and in-hospital phase after out-of-hospital cardiac arrest. Resuscitation. 2020;153:58–64. doi: https://doi.org/10.1016/j.resuscitation.2020.05.035

27. Hirlekar G, Jonsson M, Karlsson T, Hollenberg J, Albertsson P, Herlitz J. Comorbidity and survival in out-of-hospital cardiac arrest. Resuscitation. 2018;133:118–123. doi: https://doi.org/10.1016/j.resuscitation.2018.10.006

28. Andrew E, Nehme Z, Lijovic M, Bernard S, Smith K. Outcomes following out-of-hospital cardiac arrest with an initial cardiac rhythm of asystole or pulseless electrical activity in Victoria, Australia. Resuscitation. 2014;85:1633–1639. doi: 10.1016/j.resuscitation.2014.07.015

29. Mader TJ, Nathanson BH, Millay S, Coute RA, Clapp M, McNally B. Out-of-hospital cardiac arrest outcomes stratified by rhythm analysis. Resuscitation. 2012;83:1358–1362. doi: https://doi.org/10.1016/j.resuscitation.2012.03.033

30. Funada A, Goto Y, Tada H, Teramoto R, Shimojima M, Hayashi K, Kawashiri MA, Yamagishi M. Duration of cardiopulmonary resuscitation in patients without prehospital return of spontaneous circulation after out-of-hospital cardiac arrest: Results from a severity stratification analysis. Resuscitation. 2018;124:69–75. doi: 10.1016/j.resuscitation.2018.01.008

31. Reynolds JC, Frisch A, Rittenberger JC, Callaway CW. Duration of resuscitation efforts and functional outcome after out-of-hospital cardiac arrest: when should we change to novel therapies? Circulation. 2013;128:2488–2494. doi: 10.1161/circulationaha.113.002408

32. Hwang SO, Zhao PG, Choi HJ, Park KH, Cha KC, Park SM, Kim SC, Kim H, Lee KH. Compression of the left ventricular outflow tract during cardiopulmonary resuscitation. Acad Emerg Med. 2009;16:928–933. doi: 10.1111/j.1553-2712.2009.00497.x

33. Cha KC, Kim HJ, Shin HJ, Kim H, Lee KH, Hwang SO. Hemodynamic Effect of External Chest Compressions at the Lower End of the Sternum in Cardiac Arrest Patients. The Journal of Emergency Medicine. 2013;44:691–697. doi: https://doi.org/10.1016/j.jemermed.2012.09.026

34. Qvigstad E, Kramer-Johansen J, Tømte Ø, Skålhegg T, Sørensen Ø, Sunde K, Olasveengen TM. Clinical pilot study of different hand positions during manual chest compressions monitored with capnography. Resuscitation. 2013;84:1203–1207. doi: https://doi.org/10.1016/j.resuscitation.2013.03.010

35. Lah K, Križmarić M, Grmec Š. The dynamic pattern of end-tidal carbon dioxide during cardiopulmonary resuscitation: difference between asphyxial cardiac arrest and ventricular fibrillation/pulseless ventricular tachycardia cardiac arrest. Critical Care. 2011;15:R13. doi: 10.1186/cc9417

36. Grmec Š, Lah K. Difference in end-tidal carbon dioxide changes during cardiopulmonary resuscitation between cardiac arrest due to asphyxia and VF/VT cardiac arrest. Critical Care. 2003;7:P063. doi: 10.1186/cc1952

37. Lesimple A, Fritz C, Hutin A, Charbonney E, Savary D, Delisle S, Ouellet P, Bronchti G, Lidouren F, Piraino T, et al. A novel capnogram analysis to guide ventilation during cardiopulmonary resuscitation: clinical and experimental observations. Critical Care. 2022;26:287. doi: 10.1186/s13054-022-04156-0

38. Grieco DL, L JB, Drouet A, Telias I, Delisle S, Bronchti G, Ricard C, Rigollot M, Badat B, Ouellet P, et al. Intrathoracic airway closure impacts CO(2) signal and delivered ventilation during cardiopulmonary resuscitation. Am J Respir Crit Care Med. 2019;199:728–737. doi: 10.1164/rccm.201806-1111OC

39. Gutiérrez JJ, Sandoval CL, Leturiondo M, Russell JK, Redondo K, Daya MR, Ruiz de Gauna S. Contribution of chest compressions to end-tidal carbon dioxide levels generated during out-of-hospital cardiopulmonary resuscitation. Resuscitation. 2022;179:225–232. doi: https://doi.org/10.1016/j.resuscitation.2022.07.009

40. Leturiondo M, Ruiz de Gauna S, Gutiérrez JJ, Alonso D, Corcuera C, Urtusagasti JF, González-Otero DM, Russell JK, Daya MR, Ruiz JM. Chest compressions induce errors in end-tidal carbon dioxide measurement. Resuscitation. 2020;153:195–201. doi: 10.1016/j.resuscitation.2020.05.029

41. Anderson KL, Fiala KC, Castaneda MG, Boudreau SM, Araña AA, Bebarta VS. Left ventricular compressions improve return of spontaneous circulation and hemodynamics in a swine model of traumatic cardiopulmonary arrest. J Trauma Acute Care Surg. 2018;85:303–310. doi: 10.1097/ta.0000000000001901

42. Jung WJ, Cha KC, Kim YW, Kim YS, Roh YI, Kim SJ, Kim HS, Hwang SO. Intra-arrest transoesophageal echocardiographic findings and resuscitation outcomes. Resuscitation. 2020;154:31–37. doi: 10.1016/j.resuscitation.2020.06.035

